# Interpretable Deep Learning for Improving Cancer Patient Survival Based on Personal Transcriptomes

**DOI:** 10.1101/2022.10.19.22281279

**Authors:** Bo Sun, Liang Chen

## Abstract

Precision medicine chooses the optimal drug for a patient by considering individual differences. With the tremendous amount of data accumulated for cancers, we develop an interpretable neural network to predict cancer patient survival based on drug prescriptions and personal transcriptomes (CancerIDP). The deep learning model achieves 96% classification accuracy in distinguishing short-lived from long-lived patients. The Pearson correlation between predicted and actual months-to-death values is as high as 0.937. About 27.4% of patients may survive longer with an alternative medicine chosen by our deep learning model. The median survival time of all patients can increase by 3.9 months. Our interpretable neural network model reveals the most discriminating pathways in the decision-making process, which will further facilitate mechanistic studies of drug development for cancers.

## Introduction

Deep learning techniques take many forms and have shown great promise in various fields^1–5^. There has been significant interest in applying deep learning to advance biological or biomedical sciences. AlphaFold^6^ computationally predicts the protein structure at an unprecedented accuracy with an average error of approximately 1.6 angstroms. DCell^7^ predicts the impact of genetic mutations on cellular growth response. DrugCell^8^ embeds chemical structures of drugs into neural networks to predict the drug response of human cancer cell lines based on DNA point mutations. Besides their applications to cancer prognostics and therapeutics^9,10^, deep neural networks (DNN) have also been applied to cancer survival analysis^11,12^, focusing on integrating multimodal data^13^ and interpretability^14,15^. However, no deep learning method has yet considered drug treatment information in survival prediction. By incorporating the relationship between drug treatment, transcriptome, and survival, we can advance the goal of delivering personalized medicine and improving cancer prognosis.

Patients diagnosed with the same cancer and with the same driver mutations frequently show distinct clinical features and rarely have identical responses to treatments. Genetic, transcriptomic, and other clinicopathological parameters may affect patients’ survival. For some tumor types, the most significant contribution was reported from the transcriptome^16^. How genotypes and phenotypes are intertwined in cancer clinicopathology remains unclear, but the effect of genetic and cytogenetic alterations ought to be reflected in gene expression. At the same time, environmental impacts are also reflected in transcriptomes^17,18^. With deep learning techniques, global gene expression profiling appears to be the most powerful predictor of clinical outcomes. Unlike DrugCell predicting relative cell growth from point mutations, we moved beyond the cell lines and aimed to predict the clinical survival time directly based on patients’ gene expression profiles and the chemical structures of their administered drugs. Our deep-learning model enables the optimal drug choice based on individual transcriptome data.

In this work, we performed interpretable deep learning on cancer data from The Cancer Genome Atlas (TCGA)^19^ involving 33 primary tumor types. Specifically, we obtained the gene expression of primary tumor patients from TCGA and their clinical records logged in the Genomic Data Commons (GDC) database, such as vital status, survival time, and prescribed clinical drugs. We developed a DNN and named it CancerIDP (<u>Cancer I</u>nterpretable neural network based on <u>D</u>rug prescriptions and <u>P</u>ersonal transcriptomes). The system was trained to embed drug chemical structures and gene expression profiling in the feature space and then to predict the survival time based on those embeddings. The network architecture of CancerIDP is based on the Gene Ontology (GO) structure in human cells, similar to DrugCell.

Our model is highly predictive regarding cancer patient survival: the prediction is accurate for the hold-out testing data. We report a 96% accuracy for binary classification of short-lived or long-lived patients. The Pearson’s R (i.e., Pearson correlation coefficient) is 0.937 between survival time prediction and the actual clinical record. More importantly, the separate transcriptome encoder and drug encoder empower an in-silico optimal drug selection. Through a computational brute-force search, each patient is paired with every drug to test whether an alternative drug other than the actual prescribed one works better concerning survival time. In summary, our interpretable deep learning model enables precision medicine and has great potential to improve patient health.

## Results

### TCGA cancer genomics and clinical data

To predict the survival time of cancer patients based on their transcriptome profiles and drug treatments, we obtained the transcriptome and clinical data from TCGA. A total of 4311 patients have both transcriptome data and clinical information such as drug prescriptions and survival time, covering 178 drugs and 33 cancer types. The most abundant samples are from breast and ovarian tumors (Figure 1a), and the gender distribution is 63.4% females and 36.6% males (Figure 1b). Based on the vital status (Figure 1c), 1645 patients (38.2%) were deceased. The distribution of their days-to-death is shown in Figure 1d. For the other 2666 (61.8%) alive or censored patients, we used the days-to-the-last-follow-up (distribution shown in Figure 1e) to infer whether a patient was long-lived. We trained a Gene Ontology (GO)-guided deep learning network to predict the impact of different drugs on the survival time given the patient’s gene expression profile. The actual number of patients used in our network training was slightly less, depending on whether the Morgan fingerprint of the drug is available.

**Figure 1.**
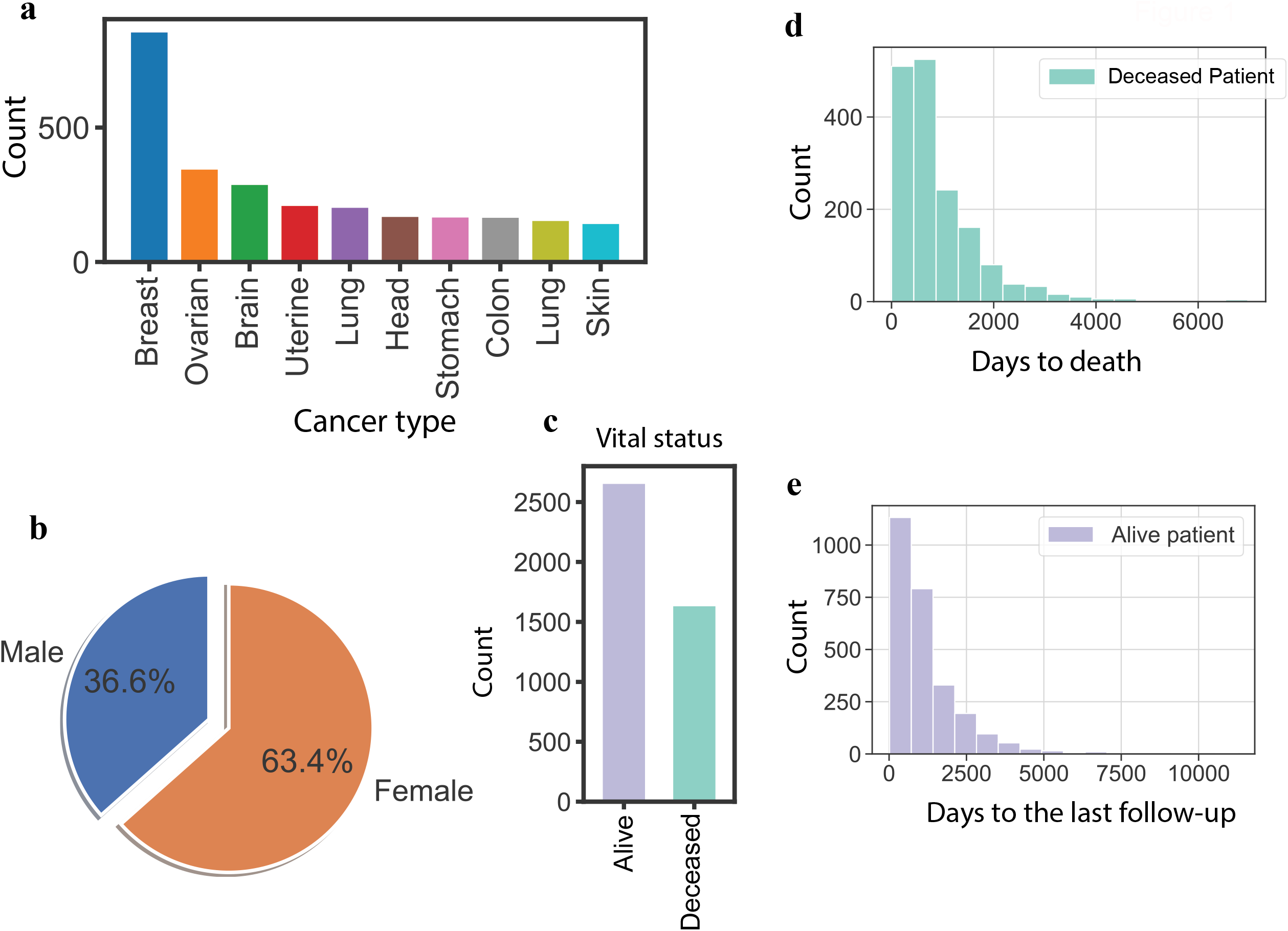
Overview of the TCGA data used in this study. (a) Top 10 cancer types from the TCGA cancer genomics data sorted by the number of patients. (b) Gender distribution. (c) Vital status from related clinical data. (d) Distribution of days-to-death for deceased patients. (e) Distribution of days-to-the-last-follow-up for alive and censored patients.

### Two-stage training with knowledge transfer to fully utilized data

Transfer learning was initially devised to transfer the learner trained in similar but different domains to the target domain^20^. Recent studies show that two-stage training can improve deep learning models in the data-scarce setting^21,22^. Here the majority of available TCGA data are censored. Therefore, to harness the power of machine learning in personalized medicine, we devised a two-stage training schema in our CancerIDP with knowledge transfer.

In the first stage, the network weights are randomly initialized, and we aim to utilize as many patients as training samples as possible. To do that, we included both deceased and qualified censored patients to create binary classification supervision signals: long-lived or short-lived. Positive samples are long-lived patients known to be alive after 1,200 days, including patients with either days-to-death or days-to-the-last-follow-up greater than 1,200. Negative samples are short-lived patients who died within 1000 days (i.e., days-to-death <1000). The first stage’s goal is to exploit as much data as possible to abstract deep representations from gene expression and drug structures for the vital status prediction (more straightforward binary classification), which will be used as a starting point in the second stage for fine-grind survival time prediction.

In the second stage, the network structure remains the same (Figure 2), and the weights are initialized as those in the best-performing model on the validation set from the first stage. The assumption is that the feature extractor learned in the binary survival status classification can be a good starting point for a more fine-grind survival time prediction. Only deceased patients with exact days-to-death records are used in the second stage. The binary network head from the first stage is discarded, and a new regression head is trained from scratch to predict the days-to-death given a patient’s gene expression profile and the prescribed drug. In the actual training, the days-to-death was transformed to months-to-death on a log scale. Figure 3a shows that the best-performing model achieves 96% classification accuracy in the first stage on the hold-out data. In the second stage, the Pearson’s R equals 0.937 between the ground truth months-to-death and the prediction (Figure 3b).

**Figure 2.**
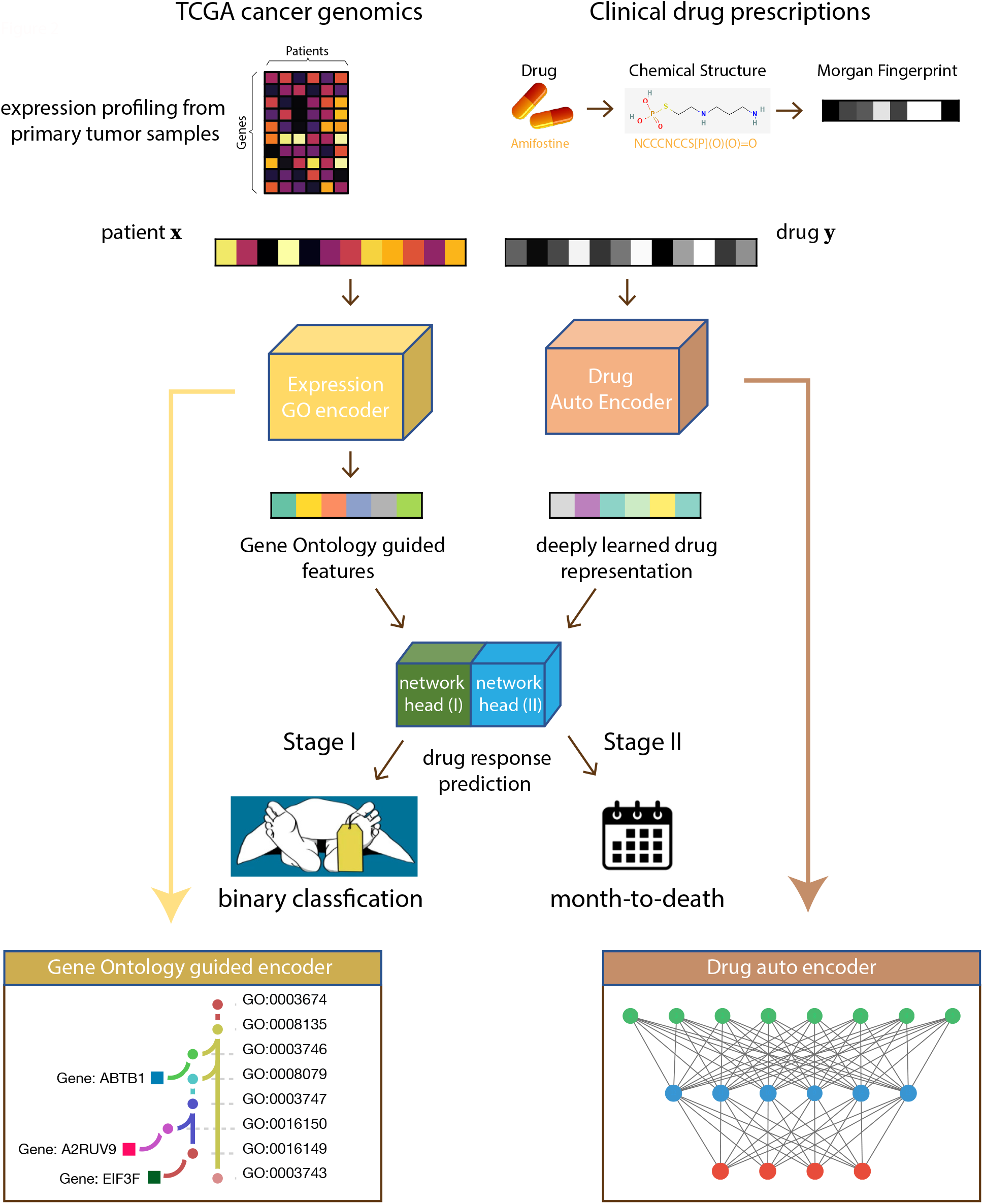
Interpretable network architecture design. Given a patient and the prescribed drug, the Gene Ontology-guided encoder takes the transcriptome as input to generate expression embedding, and the drug encoder transforms the drug Morgan fingerprints to drug embedding. The network heads then concatenate the transcriptome and drug embeddings to predict the vital status in the first stage and the exact months-to-death in the second stage. The GO-guided expression encoder follows the same GO hierarchy in human cells.

**Figure 3.**
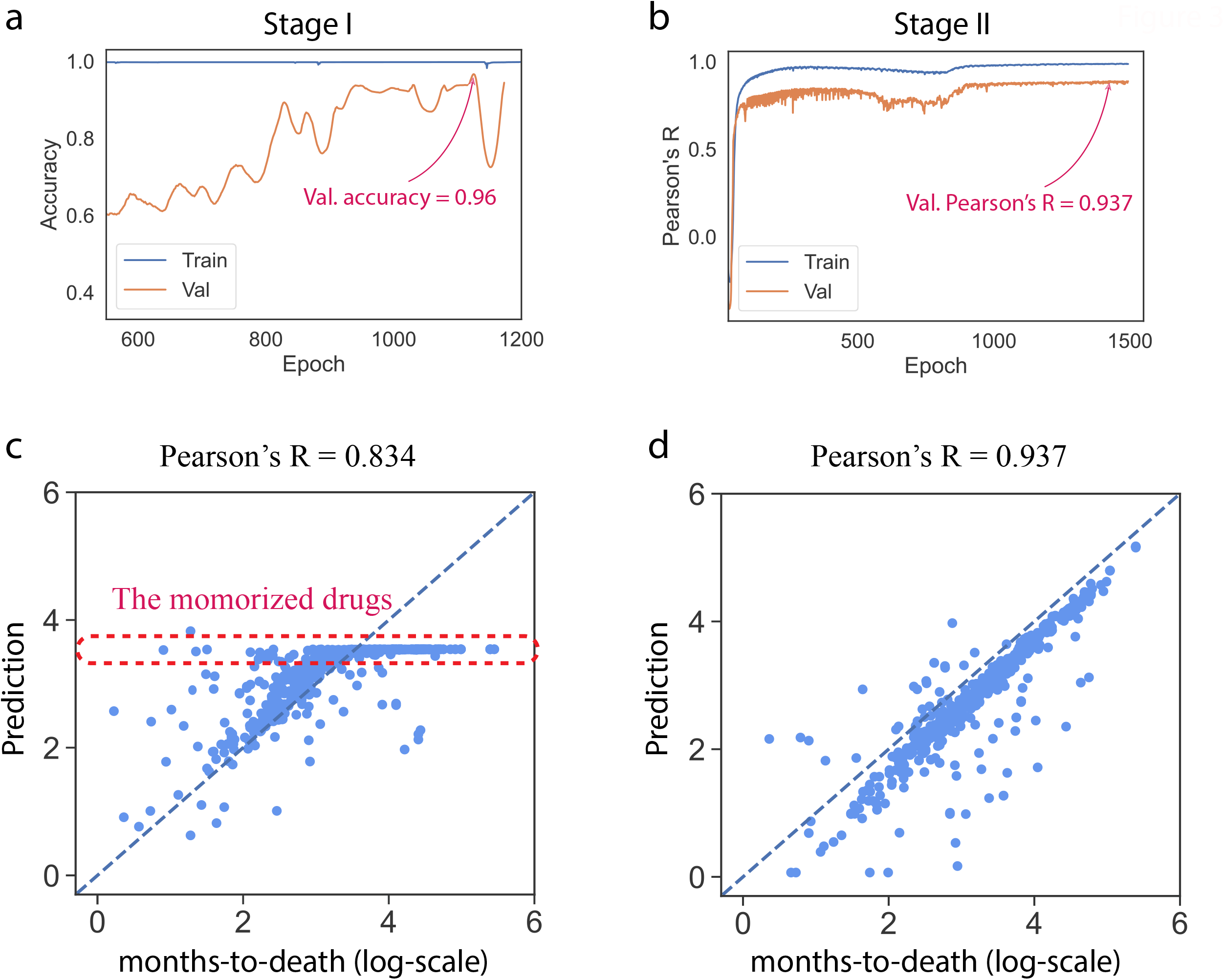
Performance evaluation and the memorization effect of the network. (a) The training and evaluation performance on hold-out validation data for vital status prediction (stage I). (b) The training and evaluation performance for survival time prediction (stage II). (c) The memorization effect for rarely prescribed drugs. (D) The performance improves after eradicating the memorized rare drugs. The Pearson’s R increases to 0.937.

### The memorization pitfall when predicting survival responses from drug treatments

Deep learning models have impressive expressiveness, but in practice, we noticed a potential pitfall that comes with the ability of the deep network to emulate any function. We called this the “memorization effect” in our survival prediction. In our earlier experiments, we included all drugs in the deep learning model and prepared the training and the hold-out validation sets by the random split. However, we observed that for some rarely prescribed drugs with only one or two supporting data points (i.e., 1-2 patients) in the training set, the network tends to simply “memorize” such drugs instead of generalizing meaningful embedding. And then, in the testing phase, the network just outputs the survival time of the patients that it has memorized (observed) in the training phase. Such effect is demonstrated in Figure 3c: some of the predicted months-to-death values (log sale) are the same since the model only observed one patient with this specific drug during the training, and the model output the survival time of that patient in the testing when fed into this particular drug.

To counter the memorization effect, we eliminated drugs with less than five supporting data points in the training set. Although the number of training samples decreases slightly, the network does not suffer from the memorization effect but learns meaningful embeddings and is more generalizable. The Pearson’s R improves from 0.834 to 0.937 after eradicating the memorization effect in the months-to-death prediction (Figures 3c and 3d).

### Improving patients’ survival based on personalized medicine enabled by CancerIDP

Distinct from other deep-learning-based cancer survival time predictions, our model not only predicts survival time but also enables optimal drug selection for each patient based on the expression encoder and the drug encoder. From the existing clinical data, we know that patient *X* with transcriptome T is prescribed drug *D*, and the survival time is *S*. This triplet (*T, D, S*) forms a training data point for our network. With the predictive ability achieved through training, the network has learned to encode transcriptomes and drugs as predictive deep embeddings. We can then predict the drug response in silico by pairing each patient with each medication and identifying the optimal drug (*D\**) with the longest survival time for patient *X*.

In this TCGA cancer genomics dataset, based on our in-silico drug selection, we can find a drug different from the actual prescription for 27.4% of patients, with which the predicted survival time is longer than the observed survival time (Figure 4a). Figure 4b compares the actual survival time for the prescribed drugs and the predicted survival time if paired with the deeply-learned optimal drugs. Deep-learning-enabled personalized medicine can significantly improve patient survival by increasing the median of months-to-death from 27.1 to 31.0. Noticeably, 23.4% of patients can have a relative improvement in survival time by at least 5% (Figure 4c, colored in orange, the size of the mark is proportional to the relative improvement). The biggest improvement is 2.97-fold, where the months-to-death is 24.7 for the prescribed drug and 73.4 for the optimal drug suggested in-silico. Deep representation learning provides great promise because as the system is fed with more abundant data, such data-driven personalized drug prescriptions will become more valuable and trustworthy.

**Figure 4.**
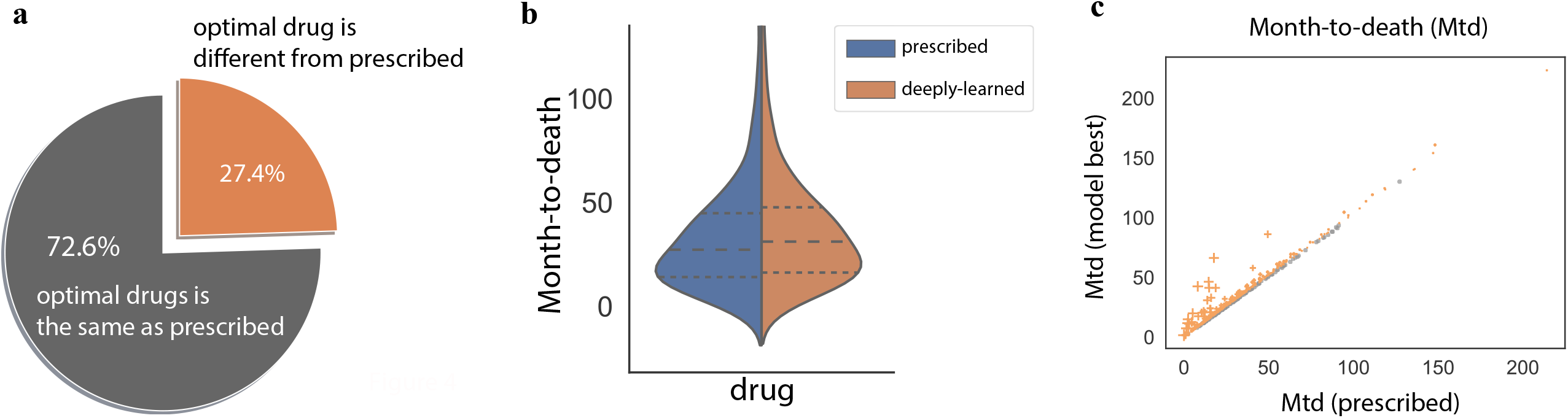
In-silico drug selection for cancer patients. (a) The percentages of patients with optimal drugs selected in-silico differ from the actually prescribed drugs. (b) The distribution of the actual months-to-death compared with the predicted months-to-death under optimal drugs selected in-silico. The dotted lines mark the 25% and 75% quantiles, and the dashed lines denote the medians. (c) The months-to-death for optimal drugs compared with the actual months-to-death. Patients with a relative improvement of >5% are colored in orange. The mark size is proportional to the relative improvement.

### Learn transcriptome embeddings that are predictive of months-to-death

The final prediction of the network is based on two components: one is the transcriptome embedding learned from the GO-guided encoder, and the other is the drug embedding from the autoencoder. The drug autoencoder is simply a black-box model that learns representations from drug chemical structures expressed in the Morgan fingerprints. We are currently unable to interpret the mechanisms. However, our transcriptome embedding is based on the hierarchy structure of GO terms and has greater interpretability of how genes act together to predict drug survival responses.

We inspected the transcriptome embedding learned from all patients using t-SNE and visualized the top two principal components (Figures 5a, 5b, and 5c). The transcriptome embedding is informative about months-to-death, as shown in Figure 5a. Patients with similar transcriptome embeddings share similar months-to-death values. The expression of some marker genes such as ESR1 (implicated in hormone resistance and anti-estrogen therapies for breast cancer, Figure 5b) or KIFC3 (encodes a member of the kinesin-14 family of microtubule motors, Figure 5c) is consistent with the spatial pattern of the transcriptome embedding, indicating the contribution of these genes to the transcriptome embedding for drug survival responses.

**Figure 5.**
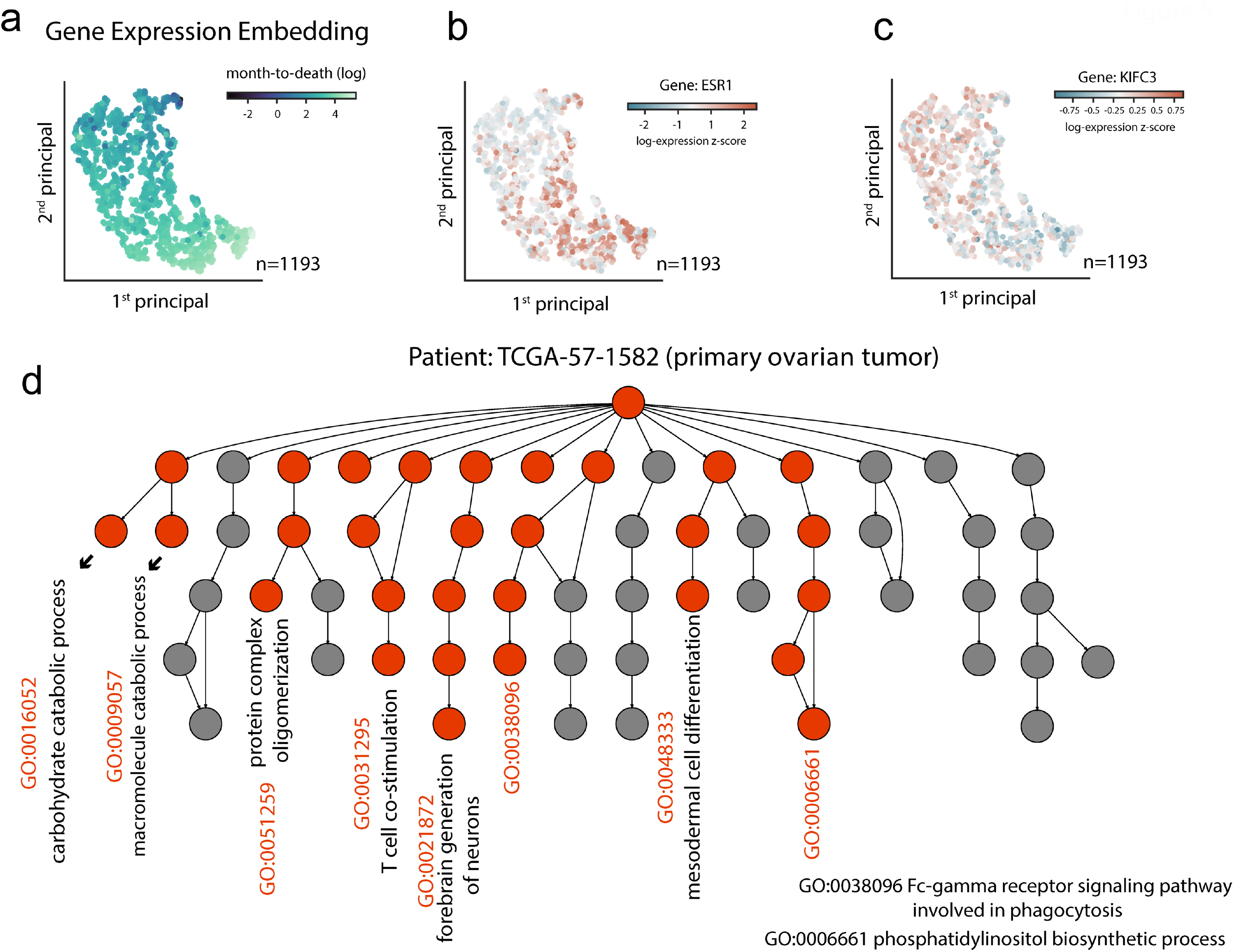
Visual inspection of gene expression embedding and the most discriminating GO entities. (a) Transcriptome embeddings learned in predicting months-to-death. The first two principal components from transcriptome t-SNE coordinates are shown. The color of the dots represents the months-to-death values. (b-c) Marker genes change along with the transcriptome embedding manifold: (b) gene ESR1; (c) gene KIFC3. The color of the dots represents expression levels. (d) The most discriminating GO terms at the individual transcriptome level. This illustrates an example from a patient with ovarian tumor.

### Discover the most discriminating GO components for a cancer patient

The transcriptome encoder is guided by the network architecture designed to be identical to the GO hierarchy in human cells. Therefore, when forward passing a patient’s gene expression profile to the trained network, the most discriminating neuron nodes in the network reflect the most distinguished biological processes (GO entities) for survival determination for that patient. Figure 5d shows an example for patient TCGA-57-1582 with ovarian cancer. The most discriminating GO terms are highlighted in red, including catabolic processes like carbohydrate catabolic process (GO:0016052), macromolecule catabolic process (GO:0009057), T-cell co-stimulation (GO:0031295), and some signaling pathways involved in phagocytosis. Therefore, such a GO-guided encoder informs the activated pathway at the individual level, paving the way for mechanistic study of personalized treatment.

## Discussion

We developed an interpretable deep learning model CancerIDP on TCGA cancer genomics and clinical data. The data, covering patients diagnosed with 33 different tumor types and their treatment drugs, are publicly available from the National Cancer Institute’s (NCI) Genomic Data Commons (GDC) database. With proper training techniques, we proved that cancer survival time is highly predictable from high-dimensional transcriptomic data. A 96% accuracy can be achieved to distinguish between long- or short-lived patients. The Pearson’s correlation coefficient on exact months-to-death between predictions and actual observations is 0.937. Our transcriptome encoder was designed to simulate the GO hierarchy in human cells. Instead of considering DNA point mutations from fixed cell lines like in DrugCell, our model takes gene expression data from individuals. Transcriptomes are dynamic and heterogenous, reflecting differences in genetics, environment, and lifestyle. They capture cellular changes caused by disease progression and therapy responses, while genetic data cannot.

The benefit of the GO hierarchy in the model architecture design is its interpretability. The activating neurons point to the most discriminating GO pathways for each individual. When the system finishes training with high predicting capability, we presume the network forms an internal understanding of the relationship between patients’ survival, their transcriptomes, and drug chemical structures. We hence propose an in-silico drug selection to search for a more desirable treatment. This in-silico drug optimization, combined with the interpretability of the GO structured expression encoder, provides data-driven precision medicine guidance for clinical practice.

In our cancer survival analysis, the survival time is the time from drug treatment to patient death. Real-life databases often record incomplete information and lack the exact survival time of some patients. When the survival time is unknown for a patient who lost clinical follow-up, such an observation is referred to as a censored data point. The last available time point is known as censoring time, usually days-to-the-last-follow-up. Censoring observations, however, contain useful information for cancer survival modeling as censoring time provides a lower bound for survival time. Such characteristic is common to real-life data: more data contain coarse information than precise information. This calls for advanced models to utilize the data fully.

Recently, two-stage (pre-train + fine-tuning) approaches^23,24^ were proposed to harness the deep learning models on data with such characteristics. In general, much data with coarse labeling are gathered to train simple classifiers in the pre-train stage. And then, knowledge obtained in the pre-train stage is transferred by sharing the feature extractor weights learned from the simple classifier. Finally, the learner is fine-tuned by training samples with fine-grind labeling. In the first stage of our model, we propose a binary classification such that patients known to be deceased within 1000 days of treatment are labeled short-lived (y=0) and patients with a survival time lower bound (days-to-death or days-to-the-last-follow-up) greater than 1200 days are labeled as long-lived (y=1). In doing so, the cancer transcriptomics and clinical data of some censored patients can be incorporated into the network training. Thereby, we utilize the lower bound information contained in the censored patients to its full potential. Then in the second stage, by sharing encoder weights, we transfer the learned knowledge for transcriptome and drug encoding and train a new network head for exact survival time prediction. In the second stage, only deceased patients with exact survival time labels are included.

Previous deep-learning models for survival analysis were applied for a single or small number of cancer types and did not consider drug and transcriptome data. These include colorectal cancer survival prediction based on histologic images and other clinical variables^15^, lung cancer survival prediction based on clinicopathologic variables^11^, and prediction for ten cancer types based on histopathologic images^25^. Our work considers all 33 primary cancer tumor types. This raises the question of whether the tissue type should be distinguished before training since tumor tissue type can be predictive to some extent for cancer survival^26^. We compared and found that when incorporating additional clinical information of the biopsies (sex, ethnicity, tissue of origin, and age at diagnosis) as additional features, the performance is on par with gene expression and drug embedding only. This indicates that the transcriptomic heterogeneity provides enough prognostic information for deep-learning-based survival prediction and that the embedding learned from high-dimensional expression data embraces human-interpretable information, at least for common features such as genders and tissues. This makes our model innately simple: it requires no extra feature engineering or data processing efforts and favors end-to-end training. The only preprocessing we performed was a cross-sample normalization for the expression matrix. The end-to-end system also eliminates potential problems for clinical data since many patient data can be missing, and how to impute missing values in a deep-learning system post significant challenges for network training^27,28^.

In summary, our CancerIDP demonstrates the great potential of deep learning models to improve cancer patient survival based on personal transcriptomes in precision medicine. With unprecedented amounts of data available in the near future, deep learning models will further enhance the predictive performance and derive essential values from the data to benefit health through such personalized approaches.

## Method

### Clinical data

Both clinical data and transcriptome profiling RNA-seq data for TCGA were obtained from the GDC data portal (https://portal.gdc.cancer.gov/repository). We obtained data for 8622 patients covering 33 cancer types and 178 unique drugs. For each patient, the drug data were obtained from the “BCR biotab” files from “clinical supplement and files.” The drug name is standardized with the drug name standardization database provided by GDISC^29^. Each drug is firstly converted to a simplified molecular-input line-entry system (SMILES) representation of its chemical structure via PubChem (https://pubchem.ncbi.nlm.nih.gov) and then encoded as a numerical vector with a fixed length (n=2048) of the Morgan fingerprint. The conversion of the SMILES representation to the Morgan fingerprint of drugs was performed with the open-source Python library RDKit. For a patient treated with different drugs at different time points or a combination of medications at the same time (103 patients), the patient is paired to each drug and treated as separate data points.

### Expression data

Transcriptome profiling via RNA-seq was used as expression data. We used Fragments Per Kilobase of transcript per Million mapped reads (FPKM) expression measurements for samples with “Sample Type” being “Primary tumor.” We focus on the 3008 genes identified from the top 15% most frequently mutated genes in Cancer Cell Line Encyclopedia (CCLE)^30^ that were annotated to Gene Ontology (GO)^31^ terms. This list of genes is also referred to as “DrugCell genes”^8^. Input normalization is extensively used in deep learning models to facilitate gradient learning and speed up convergence. Compared to raw FPKM values, standardized FPKM values across individuals (subtracting its mean and dividing by its standard deviation) speed up the training and convergence and improve the final performance by 1%. The network we described in the main text was trained with the standardized FPKM.

### Survival data

Based on the vital status of the patients, we designed two experiments. The first was designed as a binary classification, with patients known to be deceased within 1000 days as short-lived (y=0) and patients known to be alive after 1200 days as long-lived (y=1). A total of 2399 patients were included (1121 short-lived patients and 1278 long-lived patients). The second experiment is a more fine-grind model, wherein we used all deceased patients (a total of 1631 patients) to train the model to predict the clinical months-to-death data recorded for each patient.

### Neural network CancerIDP construction

The neural network is composed of two branches. The transcriptome GO encoder takes the log-transformed expression values as input and produces expression embedding. The drug autoencoder takes the Morgan fingerprints of drugs and learns the deep representation of drugs. The transcriptome GO encoder is built by mimicking the directed acyclic graph (DAG) formed by GO terms with the 3008 cancer genes as leaf nodes. The GO term architecture is based on DrugCell^8^. From the leaf nodes (gene expression values) to the root formed by the GO hierarchy, each internal node (GO term) is modeled with k (k=6) neurons through a linear layer and a batch normalization layer^32^. And the input to the linear layer is the concatenation of all child states of the internal nodes. The drug autoencoder has no prior information injected and is realized by a multilayer perceptron with three layers and the number of neurons for each layer is 64, 32, and 4. Finally, the network head combines the gene expression embedding and deeply-learned drug representation to predict the binary long- or short-lived status or the months-to-death via a non-linear classifier.

### Identification of the most discriminating GO terms

The gene expression encoder is structured according to the GO hierarchy in human cells. The internal state of each term is finally brought down to a single neuron value with a neuron activation ReLU^33^. We observed that the ReLU activation values at the same depth on the GO hierarchy have the same scale while values from different depths have slightly different scales, especially for the root GO term and the sub-root GO terms. Therefore, we scaled the ReLU activation for each GO term by its depth’s mean scale. Then the most discriminating GO processes were selected by identifying network nodes with the highest relative ReLU activation values.

## Data Availability

All data analyzed in this study are described in the article. The source code is available at
https://github.com/bsun0802/CancerIDP.

## Data availability

All data analyzed in this study are described in the article. The source code is available at https://github.com/bsun0802/CancerIDP.

## Authors’ contributions

BS and LC conceived and designed the work. BS conducted formal analyses. LC supervised the study. BS and LC wrote the manuscript. All authors read and approved the final manuscript.

## References

1. He, K., Zhang, X., Ren, S. & Sun, J. Delving Deep into Rectifiers: Surpassing Human-Level Performance on ImageNet Classification. in 1026–1034 (2015).

2. Ren, S., He, K., Girshick, R. & Sun, J. Faster R-CNN: Towards Real-Time Object Detection with Region Proposal Networks. in Advances in Neural Information Processing Systems vol. 28 (Curran Associates, Inc., 2015).

3. Sun, B., Li, B., Cai, S., Yuan, Y. & Zhang, C. FSCE: Few-Shot Object Detection via Contrastive Proposal Encoding. in 7352–7362 (2021).

4. Sun, B., Yan, J., Zhou, X. & Zheng, Y. Tuning IR-Cut Filter for Illumination-Aware Spectral Reconstruction From RGB. in 84–93 (2021).

5. Liu, Z. et al. Swin Transformer: Hierarchical Vision Transformer Using Shifted Windows. in 10012–10022 (2021).

6. Jumper, J. et al. Highly accurate protein structure prediction with AlphaFold. Nature 596, 583–589 (2021).

7. Guo, C. et al. Dcell: a scalable and fault-tolerant network structure for data centers. in Proceedings of the ACM SIGCOMM 2008 conference on Data communication 75–86 (Association for Computing Machinery, 2008). doi:10.1145/1402958.1402968.

8. Kuenzi, B. M. et al. Predicting Drug Response and Synergy Using a Deep Learning Model of Human Cancer Cells. Cancer Cell 38, 672-684.e6 (2020).

9. Tran, K. A. et al. Deep learning in cancer diagnosis, prognosis and treatment selection. Genome Medicine 13, 152 (2021).

10. Rafique, R., Islam, S. M. R. & Kazi, J. U. Machine learning in the prediction of cancer therapy. Comput Struct Biotechnol J 19, 4003–4017 (2021).

11. Wang, J. et al. SurvNet: A Novel Deep Neural Network for Lung Cancer Survival Analysis With Missing Values. Frontiers in Oncology 10, (2021).

12. Hao, L., Kim, J., Kwon, S. & Ha, I. D. Deep Learning-Based Survival Analysis for High-Dimensional Survival Data. Mathematics 9, 1244 (2021).

13. Vale-Silva, L. A. & Rohr, K. MultiSurv: Long-term cancer survival prediction using multimodal deep learning. 2020.08.06.20169698 (2020) doi:10.1101/2020.08.06.20169698.

14. Hao, J., Kim, Y., Mallavarapu, T., Oh, J. H. & Kang, M. Interpretable deep neural network for cancer survival analysis by integrating genomic and clinical data. BMC Medical Genomics 12, 189 (2019).

15. Wulczyn, E. et al. Interpretable survival prediction for colorectal cancer using deep learning. npj Digit. Med. 4, 1–13 (2021).

16. Gerstung, M. et al. Combining gene mutation with gene expression data improves outcome prediction in myelodysplastic syndromes. Nat Commun 6, 5901 (2015).

17. Tung, J. & Gilad, Y. Social environmental effects on gene regulation. Cell Mol Life Sci 70, 4323–4339 (2013).

18. Choi, J. K. & Kim, S. C. Environmental effects on gene expression phenotype have regional biases in the human genome. Genetics 175, 1607–1613 (2007).

19. Tomczak, K., Czerwińska, P. & Wiznerowicz, M. The Cancer Genome Atlas (TCGA): an immeasurable source of knowledge. Contemp Oncol (Pozn) 19, A68–A77 (2015).

20. Zhuang, F. et al. A Comprehensive Survey on Transfer Learning. Proceedings of the IEEE 109, 43–76 (2021).

21. Wang, Y. et al. A tree ensemble-based two-stage model for advanced-stage colorectal cancer survival prediction. Information Sciences 474, 106–124 (2019).

22. Gupta, A., Thadani, K. & O’Hare, N. Effective Few-Shot Classification with Transfer Learning. in Proceedings of the 28th International Conference on Computational Linguistics 1061–1066 (International Committee on Computational Linguistics, 2020). doi:10.18653/v1/2020.coling-main.92.

23. Mendes, A. & Togelius, J. Multi-Stage Transfer Learning with an Application to Selection Process. Santiago de Compostela 8 (2020).

24. Li, H., Baucom, B. & Georgiou, P. Linking emotions to behaviors through deep transfer learning. PeerJ Comput. Sci. 6, e246 (2020).

25. Wulczyn, E. et al. Deep learning-based survival prediction for multiple cancer types using histopathology images. PLOS ONE 15, e0233678 (2020).

26. Iorio, F. et al. A Landscape of Pharmacogenomic Interactions in Cancer. Cell 166, 740–754 (2016).

27. Emmanuel, T. et al. A survey on missing data in machine learning. Journal of Big Data 8, 140 (2021).

28. Cheng, C.-Y., Tseng, W.-L., Chang, C.-F., Chang, C.-H. & Gau, S. S.-F. A Deep Learning Approach for Missing Data Imputation of Rating Scales Assessing Attention-Deficit Hyperactivity Disorder. Frontiers in Psychiatry 11, (2020).

29. Spainhour, J. C. G., Lim, J. & Qiu, P. GDISC: a web portal for integrative analysis of gene–drug interaction for survival in cancer. Bioinformatics 33, 1426–1428 (2017).

30. Barretina, J. et al. The Cancer Cell Line Encyclopedia enables predictive modelling of anticancer drug sensitivity. Nature 483, 603–607 (2012).

31. Ashburner, M. et al. Gene Ontology: tool for the unification of biology. Nat Genet 25, 25–29 (2000).

32. Ioffe, S. & Szegedy, C. Batch Normalization: Accelerating Deep Network Training by Reducing Internal Covariate Shift. in Proceedings of the 32nd International Conference on Machine Learning 448–456 (PMLR, 2015).

33. Rectified linear units improve restricted boltzmann machines | Proceedings of the 27th International Conference on International Conference on Machine Learning. https://dl.acm.org/doi/10.5555/3104322.3104425.

